# Comparative Meta-Analysis of the Gut Microbiome Between ADHD and Healthy Human Cohorts

**DOI:** 10.64898/2026.09.29.26364326

**Authors:** Shivangini Singh, Anika Bokey, Ashish Pathak, Ashvini Chauhan

## Abstract

Attention-deficit/hyperactivity disorder (ADHD) is a common neurodevelopmental condition characterized by inattention, impulsivity, and hyperactivity, and is believed to involve altered dopamine signaling. Growing evidence suggests that the gut microbiome may contribute to neurobehavioral conditions, including ADHD. In this study, we examined differences in gut microbiome composition, measured using metagenomics, between healthy and ADHD-affected individuals, and evaluated age- and gender-related variation within the ADHD population. We analyzed publicly available gut microbiome datasets from the American Gut Project, NCBI, and MGnify, and retrieved data from 496 ADHD and 419 healthy individuals. Further, metadata and taxonomic abundance were processed using RStudio, normalized to sequencing depth, and analyzed at the genus level. Both groups shared several gut bacteria, including *Blautia, Bacteroides, Faecalibacterium, Roseburia*, and *Alistipes*, indicating a broadly similar core microbiome. However, individuals with ADHD showed differences in the stability and persistence of certain core taxa, such as *Proteobacteria* and the unclassified genus *Prevotellaceae, which were* uniquely present in the ADHD group. Further, PERMANOVA showed that age and gender explained a small yet significant difference in microbial variation (R^2^ = 0.0437, p = 0.001), with greater variability observed between males and females than across age groups. Dispersion analysis revealed significant microbial variability between males and females (p = 0.001), whereas age groups showed no significant variation (p = 0.792). Overall, these results suggest that differences in the gut microbiome between individuals with ADHD and healthy individuals are subtle. Clinically, this highlights the need to consider broader biological, environmental, and lifestyle influences-such as diet, medication use, and comorbidities-when evaluating the potential role of the gut microbiomes in ADHD.

**Graphical Abstract:** 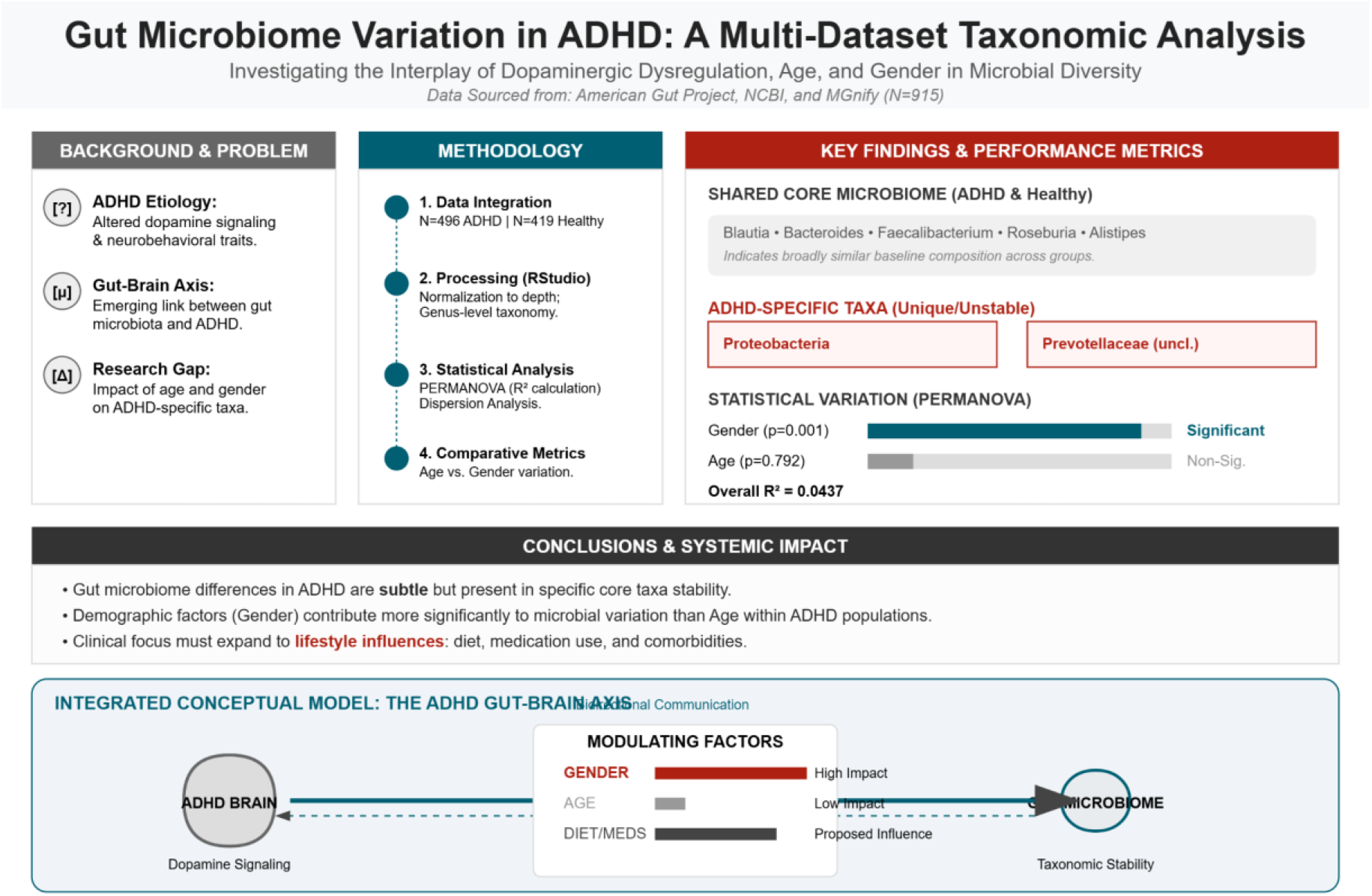

## 1. Introduction

Attention-Deficit/Hyperactivity Disorder (ADHD) is a common neurodevelopmental brain disorder characterized by persistent patterns of inattention, hyperactivity, and impulsivity that interfere with academic, occupational, and social functioning [1,2]. Although ADHD is often diagnosed during childhood, symptoms frequently persist in adolescence and adulthood, contributing to long-term psychological, behavioral, and functional impairments [2,3]. The disorder is highly heterogeneous, with considerable variation in symptom presentation, severity, and associated comorbidities, including anxiety, depression, substance use disorders, and metabolic disturbances [3].

The underlying mechanisms of ADHD remain incompletely understood despite extensive research. Current evidence suggests that ADHD arises from a complex interaction between genetic susceptibility and environmental influences [4]. Alterations in neurotransmitter systems, particularly those involving dopamine and serotonin, have been widely implicated in the pathophysiology of ADHD [5]. More recently, inflammatory pathways, immune dysfunction, and neurodevelopmental alterations have emerged as important contributors to disease progression, highlighting the disorder’s multifaceted biological basis [6,7]. More recently, increasing attention has focused on the role of the gut microbiome in neurodevelopment and mental health. Human guts harbor a complex and diverse microbial ecosystem that communicates bidirectionally with the central nervous system through the gut-brain axis [8] (Fig. 1). This communication occurs through multiple interconnected pathways, including neural mechanisms such as vagal and neurotransmitter signaling, neuroendocrine processes mediated by the hypothalamic-pituitary-adrenal axis, and immune pathways involving cytokines and inflammatory mediators. In turn, the brain influences gut microbial composition and activity through autonomic nervous system regulation, stress hormone release, immune responses, and alterations in gut motility and secretion. Disruptions in these pathways may contribute to changes in brain function and increase susceptibility to neurodevelopmental disorders, including attention-deficit/hyperactivity disorder (ADHD). Through the integration of neural, immune, endocrine, and metabolic signaling networks, the gut microbiome can influence brain development, cognition, and behavior [9]. Notably, microbial metabolites, regulation of neurotransmitter synthesis and precursors, maintenance of intestinal barrier integrity, and modulation of immune and inflammatory responses have been identified as key mechanisms through which gut microorganisms affect cognitive functioning, emotional regulation, and neurodevelopment [8,10].

**Fig. 1.**
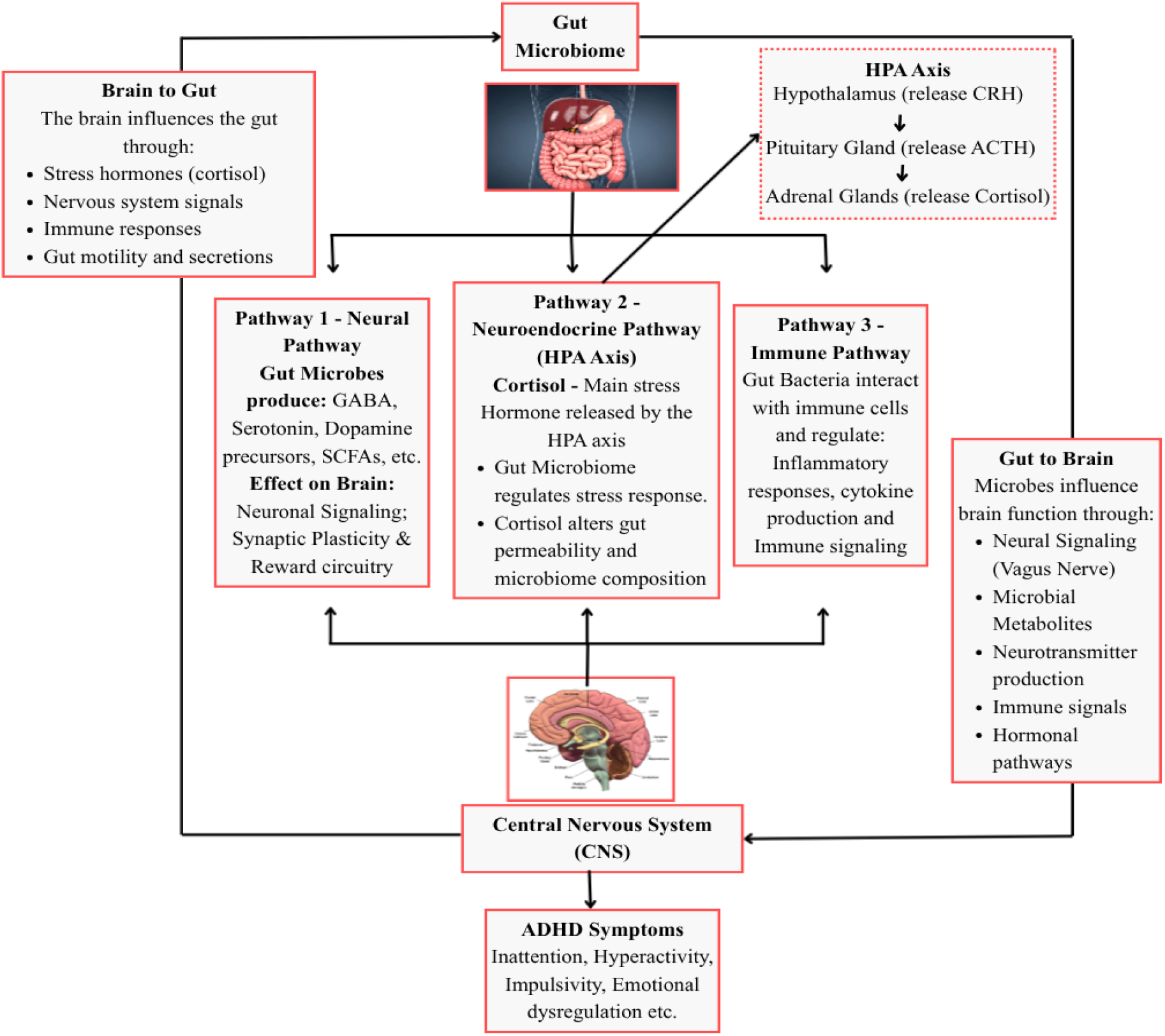
Schematic representation of the microbiota–gut–brain axis illustrating the bidirectional communication between the gut microbiome and the central nervous system. The gut microbiome communicates with the brain through neural (vagus nerve and neurotransmitter signaling), neuroendocrine (hypothalamic–pituitary–adrenal axis), and immune pathways (cytokine and inflammatory signaling). Conversely, the brain modulates gut microbial composition and function through stress hormones, autonomic nervous system activity, immune responses, and regulation of gut motility and secretion. Dysregulation of these interconnected pathways may contribute to alterations in brain function and the development of attention-deficit/hyperactivity disorder (ADHD).

Among the mechanisms linking the gut microbiome to neuropsychiatric disorders, the production of short-chain fatty acids (SCFAs), cytokine-mediated immune signaling, and vagus nerve communication are considered particularly important [8,9]. These pathways contribute to the regulation of neuroinflammation, neurotransmission, and neuronal development. Experimental and clinical studies increasingly suggest that alterations in gut microbial communities may influence behavioral outcomes and cognitive function, supporting a potential role for the microbiome in the development and progression of ADHD and other neurodevelopmental disorders [11,12].

Several investigations have examined gut microbiome composition in individuals with ADHD, reporting differences in microbial diversity and the abundance of specific bacterial taxa compared with healthy controls [13-15]. Certain microbial genera, including *Coprococcus, Prevotella*, and *Intestinibacter*, have been proposed as potential contributors to ADHD-related symptoms through their effects on neurotransmitter metabolism, inflammatory signaling, and gut-brain communication [16]. However, findings across studies remain inconsistent, and no definitive microbial signature of ADHD has yet been established [17]. While some studies have identified significant alterations in microbial composition, others have reported little or no difference in overall microbial diversity between ADHD and control populations [14,18].

Current treatment strategies for ADHD primarily include behavioral interventions and pharmacological therapies, particularly stimulant medications [19]. Behavioral approaches have demonstrated benefits in improving emotional regulation, self-management skills, and overall functioning [20]. However, pharmacological treatments may not be effective for all individuals and can be associated with adverse effects, adherence challenges, or concerns regarding long-term use [19]. Consequently, there is increasing interest in identifying novel gut microbiome-mediated biological mechanisms that may contribute to ADHD pathogenesis and offer opportunities for alternative therapeutic approaches.

Despite growing interest in the ADHD–microbiome relationship, several methodological limitations continue to hinder progress in the field. Many studies have relied on relatively small sample sizes, increasing the likelihood of false-positive findings and limiting statistical power [17]. Additionally, substantial interindividual variability in microbiome composition, together with differences in age, sex, ethnicity, diet, medication exposure, sequencing methodologies, and bioinformatic pipelines, complicates comparisons across studies [21,22]. Variations in statistical approaches used to address compositionality and zero-inflation effects further contribute to inconsistent results [23].

Given the expanding evidence linking gut microbial communities to neurodevelopmental and psychiatric conditions, further investigation is warranted to clarify the relationship between gut biota and ADHD. A more comprehensive understanding of microbiome alterations associated with ADHD may provide valuable insights into disease mechanisms, facilitate the identification of potential biomarkers, and support the development of microbiome-based interventions. Therefore, the present study aims to investigate the association between gut microbiomes and ADHD, with a particular focus on identifying microbial taxa and microbiome-related pathways that may contribute to the disorder’s pathophysiology. By employing rigorous analytical methods and accounting for relevant confounding factors, this study seeks to advance the current understanding of the gut–brain axis in ADHD and contribute to the emerging field of microbiome-based therapeutic agents, also known as psychobiotics [24].

## 2. Materials and Methods

### 2.1. Data collection and study design

A literature search was conducted to investigate differences in the gut microbiome between healthy individuals and those diagnosed with ADHD. The data acquisition for this project was done using the Mgnify database ([MGnify - EBI](https://www.ebi.ac.uk/metagenomics/)) [25] via text search - ((“human”) and (“gut”)), which resulted in a total of 150 studies. Further, Screening was conducted to identify relevant studies with ADHD datasets and healthy cohorts. The metadata for the same study was retrieved from NCBI (Run Selector: NCBI) to further screen for relevant data [26].

### 2.2. Metadata Processing

To identify the relevant dataset, metadata was manually screened for samples undergoing ADHD that also contained other diseases. To make our study more specific to ADHD, we filtered all other disease conditions to retain samples with only ADHD. To identify healthy cohorts, we excluded all disease conditions and retained samples with no disease or unhealthy symptoms. We also considered individuals’ age and gender to compare the gut microbiome across age groups in the downstream analysis, and to compare the core microbiomes between ADHD and healthy subjects.

### 2.3. Core Microbiome Analysis

After finalizing and sanitizing the datasets, we extracted the core microbiome by applying prevalence and median abundance thresholds of >= 0.5 and >= 0.001 for both datasets. This prevalence- and abundance-based approach is widely used to identify microbial taxa consistently present across samples and to define the core microbiome [27-29]. Core microbiome extraction was performed in Microsoft Excel, and the resulting core taxa were visualized as heatmaps using RStudio.

### 2.4. Alpha diversity analysis

Alpha diversity analysis was done using the Shannon diversity index, which incorporates both taxonomic richness and evenness [30,31]. Shannon diversity values were calculated for each sample from the genus-level relative abundance table. Differences in alpha diversity among the six age groups (1–20, 21–40, 41–50, 51–60, 61–70, and >70 years) were evaluated using the Kruskal–Wallis rank-sum test [32]. Comparisons between genders were performed using the Wilcoxon rank-sum test [33]. To further investigate potential sex-specific differences within individual age categories, pairwise gender comparisons were conducted separately for each age group using the Wilcoxon rank-sum test. A p-value < 0.05 was considered statistically significant.

### 2.5. Beta-dispersion analysis

The variability of the gut microbial community within each group was assessed using beta-dispersion analysis (PERMDISP) based on Bray–Curtis dissimilarity. Differences in within-group dispersion were assessed separately for age groups and sex using permutation-based analysis with 999 permutations. Differences in microbial community dispersion among age groups and between sexes were tested using a permutation test with 999 permutations [34-36]. All downstream statistical analyses and graphical visualizations were performed in **RStudio** using the **phyloseq** and **vegan** packages [37,38].

## 3. Results

### 3.1. Dataset Retrieval and Characteristics of the Study Population

Out of 150 studies identified through MGnify [25], search, only one study matched our criteria of samples with ADHD with MGnify ID “MGYS00000596” and ENA accession is “PRJEB11419”, titled “American Gut Project” (AGP) [39]. This is the largest crowdsourced citizen science project to date, providing detailed health, lifestyle, and dietary data for each sample [39]. The study consisted of 39043 samples but contained only the raw taxonomic abundance file [25]. Metadata was retrieved from NCBI [26]. Furthermore, metadata obtained from NCBI was used to screen for relevant samples. The screening process was performed completely in Microsoft Excel. The initial screening resulted in ADHD samples, which also included other diseases, such as diabetes, kidney disease, lung problems, migraine, skin condition, acne problem, thyroid, flu, IBD, chickenpox, etc. To retain “ADHD only” samples, we filtered out all the samples recorded for any other health conditions other than ADHD. This resulted in a total of 496 “ADHD only” samples. Furthermore, for healthy subjects, we excluded all unhealthy conditions, medications, and symptoms [40,41]. We obtained a total of 419 healthy samples (Fig. 2). The processed dataset underwent core microbiome analysis, alpha- and beta-diversity assessments, and PERMANOVA to evaluate differences in microbial community structure between groups. We also recorded the participants’ ages and genders, the distributions of which are shown in Fig. 3. Participants were categorized into six age groups (1–20, 21–40, 41–50, 51–60, 61–70, and >70 years). Bars represent the number of females, males, and participants with unavailable gender information (NA) within each age group for both the ADHD and healthy control cohorts.

**Fig. 2.**
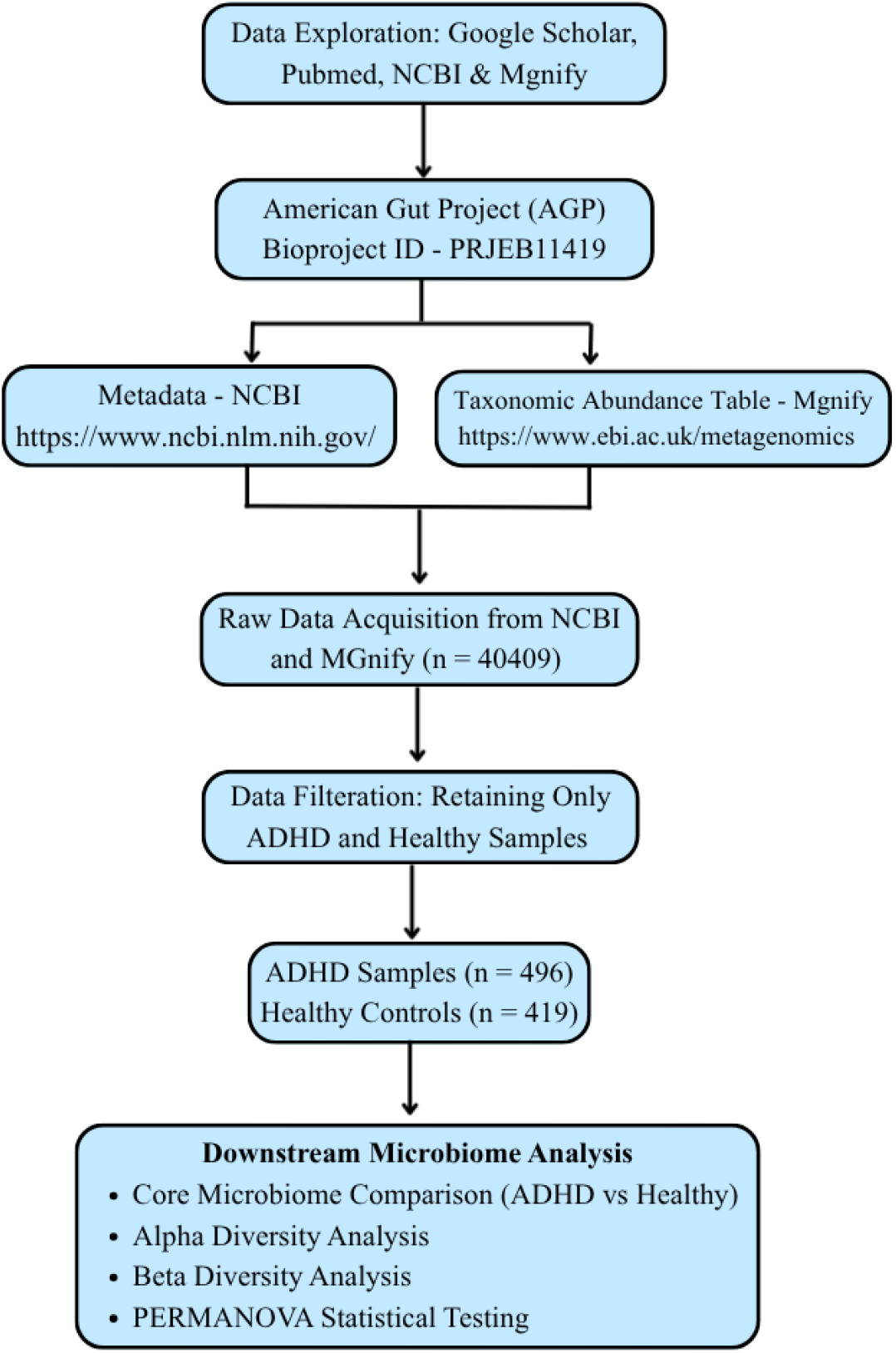
Flowchart depicting the stepwise pipeline for data screening and downstream microbiome analysis. Raw datasets obtained from public repositories were curated and filtered using Microsoft Excel and RStudio to retain ADHD and healthy control samples. The processed dataset underwent core microbiome analysis, alpha- and beta-diversity assessments, and PERMANOVA to evaluate differences in microbial community structure between groups.

**Fig. 3.**
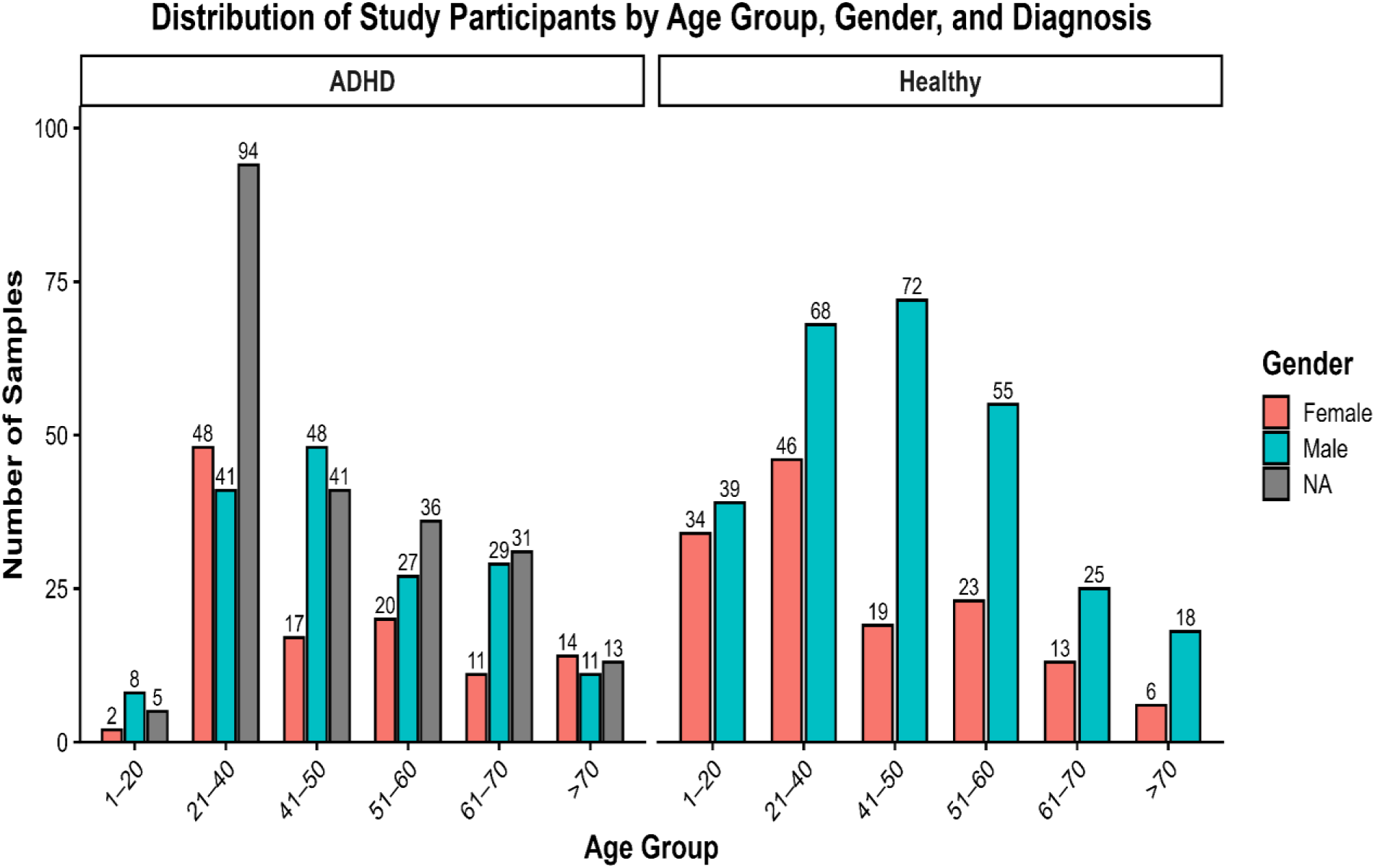
Distribution of the study participants across age groups and gender in the ADHD and healthy control cohorts following participant selection and metadata curation. Participants were categorized into six age groups (1–20, 21–40, 41–50, 51–60, 61–70, and >70 years). Bars represent the number of females, males, and participants with unavailable gender information (NA) within each age group for both the ADHD and healthy control cohorts.

After successful data acquisition and screening, the taxonomic abundance file with 39043 samples was curated based on ADHD and healthy subjects identified through metadata screening. Further, the lineage was scored to the genus level; any genus category with “NA” was replaced with a higher taxonomic level. For example, *Halobacteriaceae_ugn_19*, where the genus was not identified, so we prioritized the highest taxonomic level present, i.e., the family *Halobacteriaceae*, and “ugn_19” represented an unclassified genus in row 19. Similar hierarchical taxonomic assignments have been adopted in microbiome studies to preserve partially classified taxa for ecological analyses [42, 43]. All these filtering and sanitization were performed using Microsoft Excel in .csv format. Further, the data was used for downstream analysis.

### 3.2. Core Microbiome Analysis

Based on core microbiome analysis using the prevalence and median abundance-based thresholds of ≥ 0.5 and ≥ 0.001, different bacterial genera were identified for each group, with some common between both groups (Fig. 4). The shared core included several genera, such as *Bacteroides, Faecalibacterium, Anaerostipes, Alistipes, Parabacteroides*, and *Roseburia*, indicating the presence of a common gut microbial core irrespective of any specific health conditions like ADHD [44-48]. However, differences in the persistence of individual taxa were also observed as the detection thresholds increased. The healthy cohort retained few core genera, including *Romboutsia* and *Bifidobacterium* [49,50], whereas the ADHD cohort was characterized by the presence of unclassified genera within the families *Prevotellaceae* and *Proteobacteriota*, respectively.

**Fig. 4.**
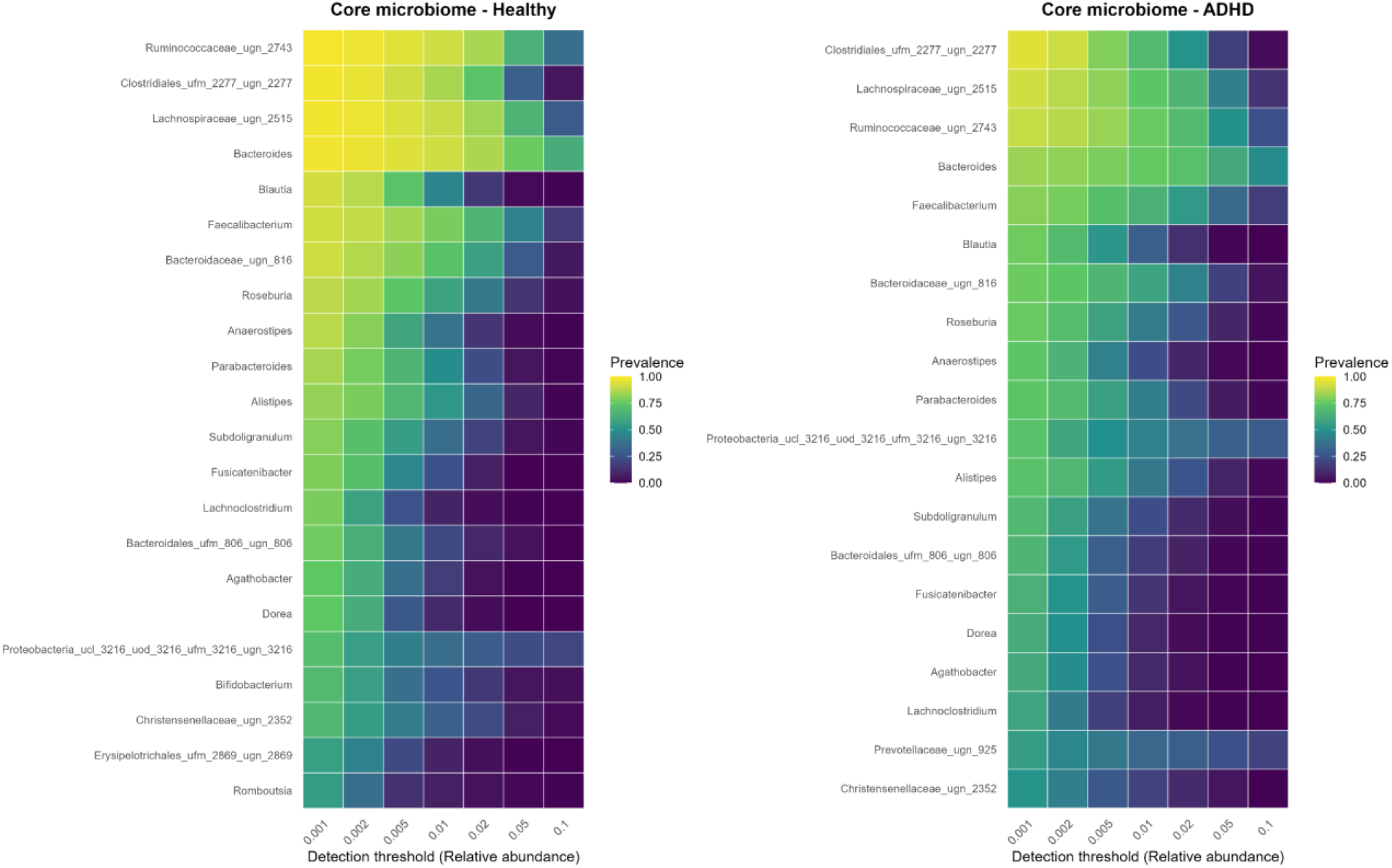
Genus-level core microbiome of ADHD and healthy cohorts identified using prevalence (≥ 0.5) and median relative abundance (≥ 0.001) thresholds. Heatmaps illustrate the distribution and persistence of core bacterial genera across increasing detection thresholds in the ADHD and healthy cohorts.

### 3.3. Alpha Diversity Analysis

The alpha diversity analysis revealed that microbial diversity did not differ significantly across the six age groups within the ADHD samples (Fig. 5a); only slight variations were observed. The overall differences were not statistically significant based on the Kruskal–Wallis test, indicating that age alone had little influence on within-sample microbial diversity. In contrast, gender showed a modest but statistically significant difference (Fig. 5b) [32]. A comparison between genders using the Wilcoxon rank-sum test revealed a significant difference in Shannon diversity (p = 0.042) [33]. While the distributions largely overlapped, male participants exhibited slightly greater variability in alpha diversity than females. To further determine whether sex-specific differences were influenced by age, Shannon diversity was compared between males and females within each age category (Fig. 5c). No significant differences were detected within any individual age group (Wilcoxon test; 1–20 years: p = 1.00; 21–40 years: p = 0.086; 41–50 years: p = 0.51; 51–60 years: p = 0.48; 61–70 years: p = 0.45; >70 years: p = 0.27). Although the 21–40-year group showed a trend toward a sex-related difference, it did not reach statistical significance.

**Fig. 5.**
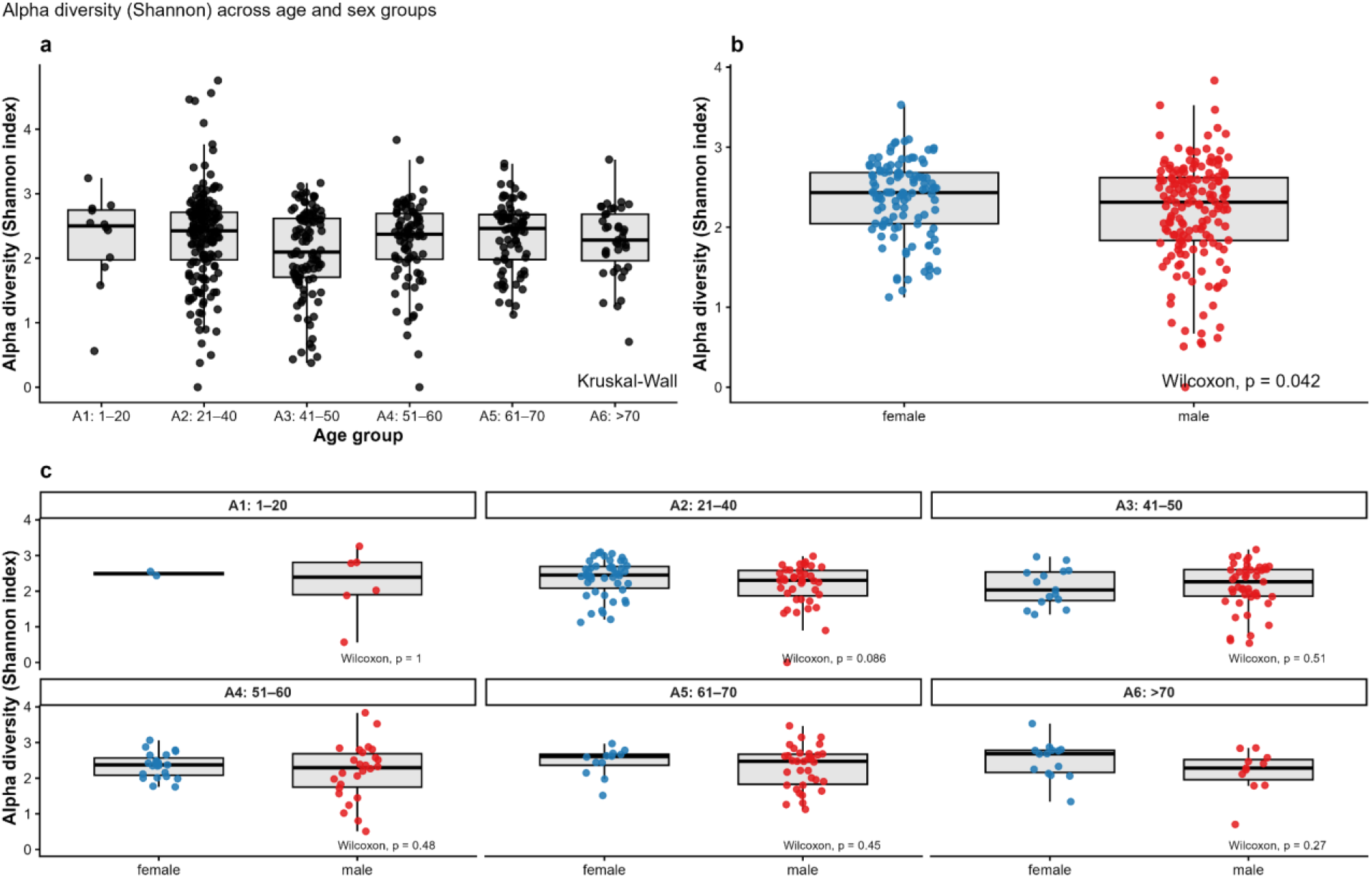
Shannon alpha diversity of the gut microbiome across age groups and sex in individuals with ADHD. (a) Comparison of Shannon diversity across six age groups (1–20, 21–40, 41–50, 51– 60, 61–70, and >70 years) using the Kruskal–Wallis test. (b) Comparison of Shannon diversity between female and male participants using the Wilcoxon rank-sum test. (c) Pairwise comparison of Shannon diversity between females and males within each age group using the Wilcoxon rank-sum test. Boxplots represent the median and interquartile range (IQR). Statistical significance was defined as *p* < 0.05.

### 3.4. Beta dispersion analysis of the gut microbial community across age and sex

To determine whether variability in gut microbial community composition differed across demographic groups, beta dispersion analysis was performed using Bray–Curtis dissimilarities (Fig. 6 a,b). Comparison of dispersion among the six age groups revealed no significant differences in within-group microbial variability (permutation test, p = 0.799; Fig. 6a). Although minor differences in median distances to group centroids and dispersion ranges were observed across age categories, the substantial overlap among groups indicates that age did not significantly influence the heterogeneity of the gut microbial community in individuals with ADHD. In contrast, sex had a significant effect on microbial community dispersion (permutation test, p = 0.001; Fig. 6b). Male participants exhibited greater distance from the group centroid and a wider distribution of Bray– Curtis dissimilarities than female participants, indicating higher inter-individual variability in gut microbial composition among males. Female samples, in comparison, displayed relatively lower dispersion, suggesting a more homogeneous microbial community structure.

**Fig. 6.**
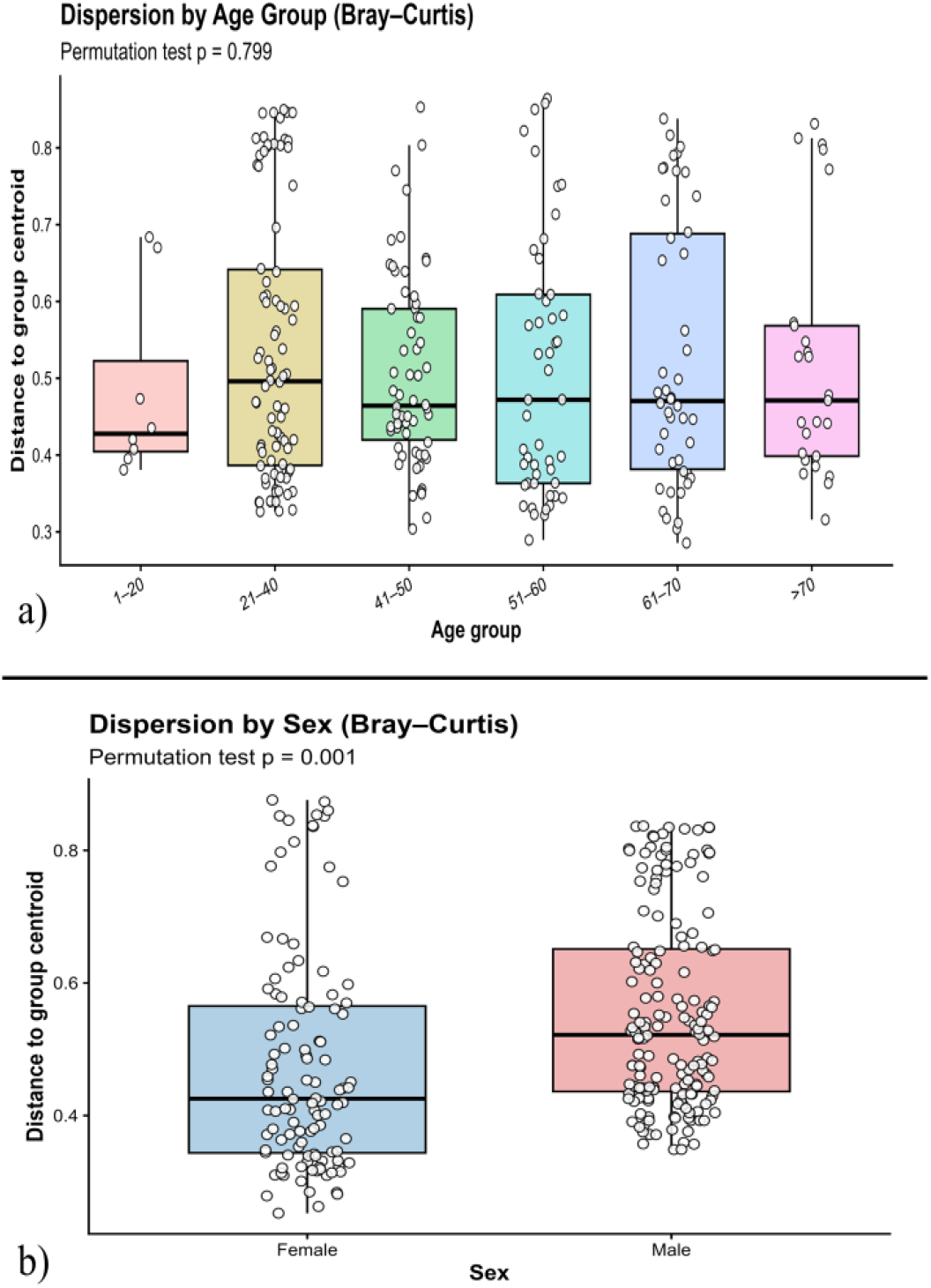
Beta-dispersion analysis of the gut microbial community across age groups and sex in individuals with ADHD based on Bray–Curtis dissimilarity. (a) Comparison of within-group microbial community dispersion among six age groups (1–20, 21–40, 41–50, 51–60, 61–70, and >70 years) using Bray–Curtis dissimilarity and a permutation test (PERMDISP; 999 permutations). No significant differences in community dispersion were observed across age groups (*p* = 0.799). (b) Comparison of microbial community dispersion between female and male participants using Bray–Curtis dissimilarity and a permutation test (PERMDISP; 999 permutations).

## 4. Discussion

This study investigated gut microbial community characteristics in individuals with ADHD using data derived from the American Gut Project, one of the largest publicly available microbiome datasets. Through rigorous metadata screening, we identified 496 individuals reporting ADHD without other comorbid conditions and compared them with 419 healthy controls. The large sample size enabled the evaluation of core microbiome composition and the influence of demographic factors, including age and sex, on microbial diversity and community structure.

### 4.1 Core Microbiome Characteristics in ADHD

Core microbiome analysis revealed substantial overlap between the ADHD and healthy control groups. Genera such as *Bacteroides, Faecalibacterium, Anaerostipes, Alistipes, Parabacteroides*, and *Roseburia* were consistently present in both cohorts, suggesting a stable microbial core maintained regardless of ADHD status. These genera are common constituents of the healthy human gut microbiome and are known contributors to short-chain fatty acid production, intestinal homeostasis, and host metabolic regulation. Their persistence in both cohorts suggests that ADHD is unlikely to be associated with large-scale disruption of dominant gut microbial taxa.

Despite this overlap, several distinctions were observed. Healthy individuals retained *Bifidobacterium* and *Romboutsia* as persistent core members at higher prevalence thresholds, whereas ADHD samples were characterized by unclassified taxa belonging to the families *Prevotellaceae* and *Proteobacteriota*. The reduced persistence of *Bifidobacterium* in ADHD is noteworthy because species within this genus have been associated with neurotransmitter precursor metabolism, immune regulation, and maintenance of gut barrier function. Alterations in *Bifidobacterium* abundance have also been reported in previous ADHD and neurodevelopmental studies, although findings have been inconsistent across populations.

The increased prevalence of unclassified *Prevotellaceae* and *Proteobacteriota*-related taxa may indicate shifts toward microbial communities with altered metabolic or inflammatory potential. Members of the phylum Proteobacteria are often considered markers of microbial instability or dysbiosis, while alterations in *Prevotella*-related taxa have previously been linked to diet, immune function, and gut-brain interactions. However, because these taxa could not be resolved to the genus level, their biological significance should be interpreted with caution.

### 4.2. Limited Influence of Age on Gut Microbial Diversity

Contrary to expectations, age did not significantly influence alpha diversity or beta dispersion within the ADHD cohort. Shannon diversity remained relatively stable across all age categories, and community heterogeneity did not differ significantly among age groups. These findings suggest that once ADHD-associated microbial patterns are established, they may remain relatively stable throughout adulthood.

Previous studies in the general population have often reported age-related shifts in microbiome composition, particularly during early childhood and advanced age. However, such effects may be less apparent in cohorts dominated by adults, as the gut microbiome is generally considered relatively stable after maturation. Furthermore, the broad age categories used in the present study may have reduced sensitivity to detect subtle age-dependent microbial changes.

The lack of age-associated differences may also reflect the influence of stronger determinants of microbiome composition, including dietary habits, medication usage, geography, and lifestyle factors, which could mask relatively small age-related effects. Therefore, while age is an important factor in microbiome development, its contribution to microbial diversity within this ADHD cohort appears limited.

### 4.3. Sex Differences in Microbial Diversity and Community Structure

A notable finding of this study was the influence of sex on variation in the gut microbiome. Although the difference in alpha diversity between males and females was relatively small, it was statistically significant. More importantly, beta dispersion analysis demonstrated substantially higher microbial community variability among males than females.

The greater dispersion observed in males suggests increased inter-individual heterogeneity in gut microbial composition. This finding may reflect differences in hormonal regulation, immune responses, dietary patterns, or lifestyle factors between sexes. Sex hormones are known to shape microbial communities by affecting host metabolism and immune function, and previous studies have reported sex-specific associations between the gut microbiota and neurodevelopmental disorders.

Interestingly, when sex differences were assessed within individual age groups, no statistically significant differences were detected. This suggests that the overall sex effect observed across the ADHD cohort may reflect cumulative differences across multiple age groups rather than pronounced effects within any single age category. The trend observed in the 21–40-year age group supports this interpretation and warrants further investigation in larger, more balanced cohorts. Taken together, these findings suggest that sex may represent a more important determinant of gut microbial variation in ADHD than age. Future ADHD microbiome studies should therefore consider stratifying analyses by sex to avoid masking biologically relevant patterns.

### 4.4. Implications for the Gut-Brain Axis in ADHD

The observed differences in core microbiome composition support growing evidence that ADHD may be associated with subtle shifts in the gut microbial ecosystem rather than dramatic losses of diversity. Microbial taxa identified in this study have been linked to short-chain fatty acid production, immune regulation, metabolic homeostasis, and neurotransmitter-related pathways, all of which contribute to gut-brain axis communication. Changes affecting these microbial functions could influence neurodevelopment, cognition, and behavior through immune, endocrine, and metabolic mechanisms.

However, the overall similarity between ADHD and healthy individuals suggests that ADHD-associated microbial alterations may be functional rather than purely taxonomic. In other words, changes in microbial metabolic activity may be more important than differences in the presence or absence of specific taxa. Future studies integrating metagenomics, metabolomics, and clinical symptom measures will be necessary to determine how microbial functions contribute to the pathophysiology of ADHD.

### 4.5. Strengths and Limitations

A major strength of this study is the use of a large, publicly available microbiome dataset, combined with stringent metadata filtering, to identify individuals who report ADHD without major comorbid conditions. The relatively large sample size provided sufficient statistical power to examine demographic influences on microbiome diversity.

Several limitations should also be considered. First, ADHD status was self-reported within the American Gut Project and could not be independently verified clinically. Second, detailed information regarding ADHD subtype, symptom severity, medication history, and duration of diagnosis was unavailable. Third, the cross-sectional design precludes causal inference regarding microbiome-ADHD relationships. Fourth, taxonomic resolution was limited to the genus level, preventing identification of species-specific associations. Finally, residual confounding from unmeasured dietary, environmental, and lifestyle factors may have influenced microbial composition.

### 4.6. Conclusion

Overall, our findings indicate that individuals with ADHD share a substantial core gut microbiome with healthy controls but exhibit differences in the persistence of specific taxa, particularly *Bifidobacterium, Romboutsia*, and unclassified members of *Prevotellaceae* and *Proteobacteriota*. Age had minimal influence on gut microbial diversity and community structure, whereas sex emerged as a significant source of microbial variation, particularly in terms of community heterogeneity. These results support the growing recognition of the gut microbiome as a potential contributor to ADHD-related biological processes while highlighting the importance of considering demographic factors, especially sex, in future microbiome research.

## Data Availability

All data produced in the present study are available upon reasonable request to the authors

## Competing interests

The author(s) declared that this work was conducted in the absence of any commercial or financial relationships that could be construed as a potential conflict of interest.

## Funding

This work was primarily supported by the U.S. Department of Defense under Award Number W911NF2310268, administered by the U.S. Army Contracting Command-Aberdeen Proving Ground-Research Triangle Park Division (ACC-APG-RTP Division). The funding supported the conduct of this research and its publication. Additionally, funding from the Department of Defense contract #W911NF2210145 and the Department of Energy (DOE) Minority Serving Institution Partnership Program (MSIPP) task order agreement #800002172 also supported this work.

### Acknowledgements

The authors gratefully acknowledge all study participants whose contributions made it possible to develop publicly available metagenomic repositories used in this research. We also thank the researchers, investigators, and institutions who generated, curated, and shared these datasets, enabling this comprehensive analysis of the human gut microbiome in a cohort diagnosed with ADHD.

## Generative AI statement

The author(s) declare that Generative AI was used in the creation of this manuscript. AI-based tools (ChatGPT, OpenAI) were used solely for language editing and formatting. The authors retain full responsibility for the content and conclusions presented in this manuscript.

## Notes

### Competing Interest Statement

The authors have declared no competing interest.

### Author Declarations

Out of 150 studies identified through MGnify, search, only one study matched our criteria of samples with ADHD with MGnify ID MGYS00000596 and ENA accession is PRJEB11419, titled American Gut Project (AGP).

